# Detection of multiple cardiac abnormalities using Convolution, Positional Encoder and Transformer on 12-lead ECG recordings

**DOI:** 10.1101/2024.06.17.24309016

**Authors:** Adam Ford, Jessica Lan

**Affiliations:** Texas Tech University, Lubbock, TX

**Keywords:** electrocardiogram, deep learning, transformer, decision support system

## Abstract

**Objectives:** Firstly, we aimed to develop a system capable of detecting multiple cardiac abnormalities simultaneously from 12-lead ECG recordings. Secondly, we tried to improve the detection by analyzing the relationship between imbalanced datasets and optimal classification thresholds.

**Methods:** A novel fusion of Convolution Positional Encoder and Transformer Encoder was used to solve the multi-label classification problem. We used a proper evaluation metric called area under the precision-recall curve (AUPRC) that enabled us to analyze the precision-recall trade-off and find the optimal thresholds.

**Results:** Having outperformed other popular deep networks, the model achieved the highest AUPRC of 0.96 and *f*_1_-score of 0.90 on 42511-sample datasets. We also found the negative correlation coefficient of -0.68 between optimal thresholds and the proportion of positive samples.

**Significance &Conclusion:** This study compared the performances of different deep learning architectures on a medical problem and showed the potential of advanced techniques in capturing spatial, temporal features alongside attention mechanisms. It also introduced how to reduce the impact of imbalanced datasets and find optimal classification thresholds.

## 1. INTRODUCTION

Cardiovascular diseases have been increasingly common in modern society nowadays, thus it is imperative to develop techniques to deal with those. One of the most effective tools to analyze and diagnose such diseases is the electrocardiogram (ECG), and making automatic diagnoses from ECG recordings has become vitally important. Traditionally, there were several techniques that used machine learning to automatically diagnose cardiovascular diseases. For example, [1–3] employed Support Vector Machine (SVM) [4] and Decision Tree [5]. However, conventional machine learning algorithms require a manual feature engineering process. It is often time-consuming and a number of potential features in the ECG signals may be neglected. To deal with this problem, modern deep learning methods have been introduced to automatically extract a far larger number of latent features in the data. For example, [6–9] used Convolutional Neural Network (CNN) [10] and Long Short Term Memory (LSTM) [11] to capture the spatial and temporal information from the signals. However, these studies assumed that the abnormalities were mutually exclusive, i.e., one recording had no more than one abnormality, whereas in practice, patients can suffer from multiple cardiac abnormalities simultaneously. In addition, a wide range of research works analyzed the performance of one deep neural network on different datasets, thus there was a lack of comparisons of multiple neural network models on the same dataset, which could give us insights into how they would perform on medical data. Besides, medical datasets are often imbalanced, i.e., the proportion of negative samples are far higher than that of positive ones, thus it is vital to use proper methods for assessment. Previous studies such as [6–8,12,13] used *f*_1_-score [14] to evaluate their models. Though *f*_1_-score can fairly take positive and negatives samples into consideration, it does not consider the whole threshold range [0, 1]. Instead, it uses the default value of 0.5: if the output probability is lower than that value, then its original signal will be classified as negative, and otherwise. There may exist an optimal threshold value that balances the precision-recall trade-off and maximizes *f*_1_-score, along-side the correlation between that optimal value and the proportion of positive samples.

Major contributions of this work are as follows:

- We developed a model that could detect multiple cardiac issues simultaneously, rather than considering mutually exclusive abnormalities.
- We trained and tested a fusion of recent advancements in Deep Learning: Residual Convolution for exploring spatial features; Positional Encoder which is able to capture temporal information in parallel, and Transformer Encoder to utilize attention mechanisms. The model outperformed other well-known architectures in detecting all abnormalities.
- We evaluated our model in the whole classification threshold range [0, 1], and found optimal threshold value for each cardiac abnormality. We also showed the negative correlation between the optimal threshold and the proportion of positive samples.

## 2. EXPLORATORY DATA ANALYSIS

We used three public datasets for the PhysioNet Challenge 2020 [15, 16], which were collected from multiple sources in three different continents. The first source was the dataset from the China Physiological Signal Challenge 2018 (CPSC2018) [17], Southeast University, China. It has a total of 10,330 recordings (5,542 males, 4,788 females), sampled at 500 Hz, with varying durations from 10 to 60 seconds. The second one was from the Physikalisch Technische Bundesanstalt (PBT-XL) [18, 19], Brunswick, Germany, with a total of 21,837 recordings (11,379 males and 10,458 females), sampled at 500 Hz for 10 seconds. The last data source was the Georgia 12-Lead ECG Challenge Database [20], collected at Emory University, Atlanta, Georgia, USA. This dataset consists of 5,551 male and 4,793 female recordings, yielding a total of 10,344. They were also sampled for 10 seconds at 500 Hz. In total, there were 42,511 recordings in our datasets.

To be compatible with deep neural networks, all data samples must have the same length. Since the CPSC2018 dataset has variable-length recordings, we either padded or truncated some recordings, to increase or reduce their length to a target one. In the three datasets, 35,948 out of 42,511 recordings last 10 seconds, amounting to more than 84%. Therefore, we chose 10-second as the target duration, corresponding to the sequence length of 10 × 500 = 5000. All recordings that were longer than 10 seconds were truncated evenly on both sides. For example, if a recording lasted 60 seconds from 0 to 59, we kept the middle 10-second sub-recording from 25 to 34 and eliminated the rest.

We trained our model to detect 13 cardiac types: a normal class (Normal Sinus Rhythm - NSR) and 12 abnormalities, namely 1st Degree AV Block (IAVB), Abnormal QRS (abQRS), Atrial Fibrillation (AF), Left Anterior Fascicular Block (LAnFB), Left Axis Deviation (LAD), Left Bundle Branch Block (LBBB), Left Ventricular Hypertrophy (LVH), Myocardial Infarction (MI), Myocardial Ischemia (MIs), Right Bundle Branch Block (RBBB), Sinus Bradycardia (SB) and Sinus Tachycardia (STach). Table 1 shows the dataset statistics.

**Table 1.** Dataset Summary.

|  |  | CPSC2018 | PBT-XL | Georgia | Total | % |
| --- | --- | --- | --- | --- | --- | --- |
| Class | IABV | 828 | 797 | 769 | 2394 | 5.63 |
|  | abQRS | 0 | 3389 | 0 | 3389 | 7.97 |
|  | AF | 1374 | 1514 | 570 | 3458 | 8.13 |
|  | RBBB | 1971 | 542 | 570 | 3083 | 7.25 |
|  | LAnFB | 0 | 1626 | 180 | 1806 | 4.25 |
|  | LAD | 0 | 5146 | 940 | 6086 | 14.32 |
|  | NSR | 922 | 18092 | 1752 | 20766 | 48.85 |
|  | LBBB | 274 | 536 | 231 | 1041 | 2.45 |
|  | LVH | 158 | 2359 | 1232 | 3749 | 8.82 |
|  | MI | 376 | 5261 | 7 | 5644 | 13.28 |
|  | MIIs | 384 | 2175 | 0 | 2559 | 6.02 |
|  | SB | 45 | 637 | 1677 | 2359 | 5.55 |
|  | STach | 303 | 826 | 1261 | 2390 | 5.62 |
| Gender | Male | 5542<br>(53.6%) | 11379<br>(52.1%) | 5551<br>(53.7%) | 22472<br>(52.9%) |  |
|  | Female | 4788<br>(46.4%) | 10458<br>(47.9%) | 4793<br>(46.3%) | 20039<br>(47.1%) |  |

## 3. METHODS

This section gives details about the proposed CNN-TE model, whose block diagram is shown in Figure 1. We describe the role of each module in the diagram as follows.

**Fig.1.**
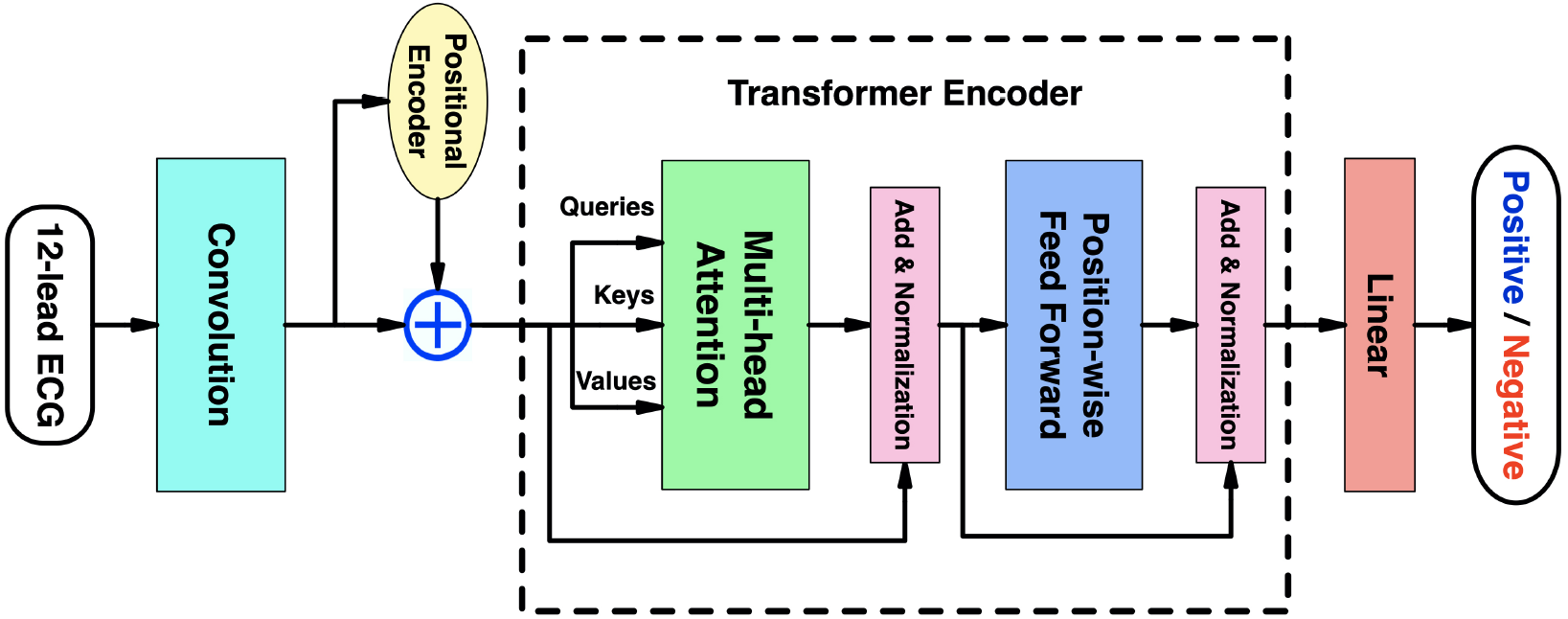
Block diagram of the CNN-TE model

### 3.1 Convolution

Convolutional Neural Networks have been widely used in Computer Vision and even Natural Language Processing applications, such as object detection, object recognition [21, 22], sentiment analysis [23]. The key components of such networks are convolutional layers, which capture spatial information and expand the feature representation of the input tensors. In other words, it reduces the spatial dimension and increases the feature dimension. Thus these layers should perform well on tasks related to processing images (2D array) or time-series signals (1D array).

In this study, we built a convolutional module which mainly consisted of 8 residual blocks [24], as illustrated in Figure 2. Given 12-lead ECG recordings as its input in the format of a tensor with shape (batch size, sequence length, num features) = (256, 5000, 12), this block reduced the sequence length (spatial dimension) from 5000 to 157, and increased the feature dimension from 12 to 512, to output a tensor with shape (256, 157, 512).

**Fig.2.**
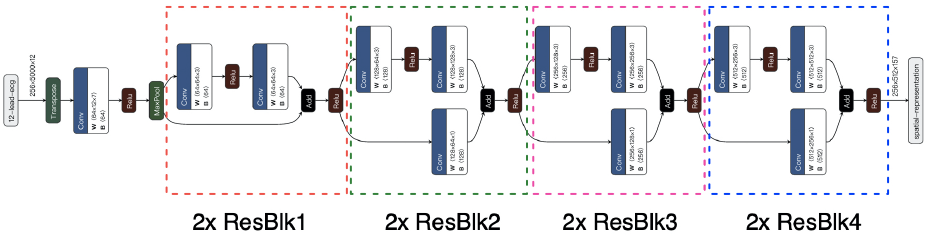
Convolutional module

### 3.2. Positional Encoder

ECG recordings can be considered to be time-series data, thus they might contain latent temporal information. It would be beneficial if such information could be captured. Recurrent Neural Networks (RNN) such as Gated Recurrent Units (GRU) [25] or Long Short Term Memory (LSTM) [11] have been widely used for that purpose. However, those recurrent models learn temporal representation from the data sequentially, i.e, they must have the result at timestep *t* −1 available to compute the result at timestep *t*, thus require a long training time. In this study, the Positional Encoder [26] was employed to inject the temporal information of the input recordings into themselves. This module is capable of capturing the information in parallel.

Let **X** ∈ **R**^*l*×*d*^ denotes an input recording of the block, where *l* is the length of the recording, *d* is the number of features at each position. The Positional Encoder encodes the position **P** ∈ **R**^*l*×*d*^ of **X** and outputs **P** + **X**. The position **P** is computed by sine and cosine functions as follows:

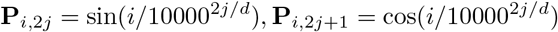

Here, *i* = 0, …, *l* − 1 represents the position of each element in the recording, *j* = 0, …, ⌊(*d*−1)*/*2⌋ refers to the feature dimension. Specifically, after the convolutional module, the recording length was *l* = 157 and the number of features was *d* = 512. Since **P**_*i*,2*j*_ and **P**_*i*,2*j*+1_ are independent of other components at *i* −1, *i* −2, …, they can be computed in parallel, resulting in faster training time. The input and output of the positional encoder were added, then passed to the Transformer Encoder.

### 3.3 The Transformer Encoder

#### 3.3.1 Multi-head Attention

Attention mechanism [27, 28], based on the idea in cognitive science that we should focus on some key parts when processing a huge amount of information, is one of the most powerful techniques in Deep Learning nowadays. Mathematically, suppose that an attention layer has a “memory”, in which past information has been stored in the form of *n* key-value pairs: (**k**_1_, **v**_1_), …, (**k**_*n*_, **v**_*n*_), with 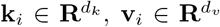. Given an input 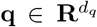, called a *query*, the attention layer returns an output that has some parts being focused more than others, based on the following procedure.

Firstly, the layer computes the similarity **s** = (*s*_1_, …, *s*_*n*_) of the input query **q** with each key **k**_1_, …, **k**_*n*_ by a scoring function *S*:

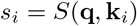

The scoring functions *S* are often either dot-product or multi-layer perceptron [29]. The Transformer architecture uses scaled dot-product attention (DPA) [26], which is defined by:

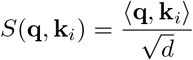

Here the query **q** and the key **k**_*i*_ have the same dimension *d*_*q*_ = *d*_*k*_ = *d*, ⟨**q, k**_*i*_⟩ is the dot-product of the two *d*-dimensional vectors **q** and **k**_*i*_. The dot-product is then scaled by a factor of 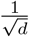 to prevent vanishing gradients when *d* is large [26].

Then, the similarities are passed through a softmax function to produce attention weights **w** = [*w*_1_, …, *w*_*n*_]^*T*^ :

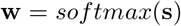

or

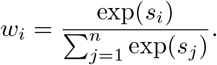

Finally, the attention layer’s output *o* is determined by the weighted sum of the values:

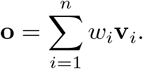

In short, the DPA can be described as:

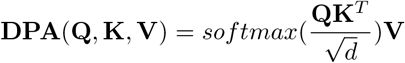

where **Q** ∈ **R**^*n*×*d*^, **K** ∈ **R**^*n*×*d*^, 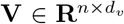 are arrays of query, key and value vectors.

The Transformer used self-attention mechanisms, in which the query, key, and value are identically taken from each element of the input sequence. Figure 3 shows the architecture of a multi-head attention layer. Instead of processing the entire *d*-dimensional queries, keys, and values, the Transformer Encoder split the feature dimension equally into *h* DPA blocks called *heads*, projects the queries, keys, and values with 3 Linear layers, performs the attention function parallelly, then concatenate the results from each head, and eventually passes the concatenated result to a final Linear layer. This mechanism enables the model to focus on different feature subspaces at different positions. Moreover, the output can be computed in parallel, thus the multi-head attention layer should be more efficient than sequential-computing recurrent neural networks.

**Fig.3.**
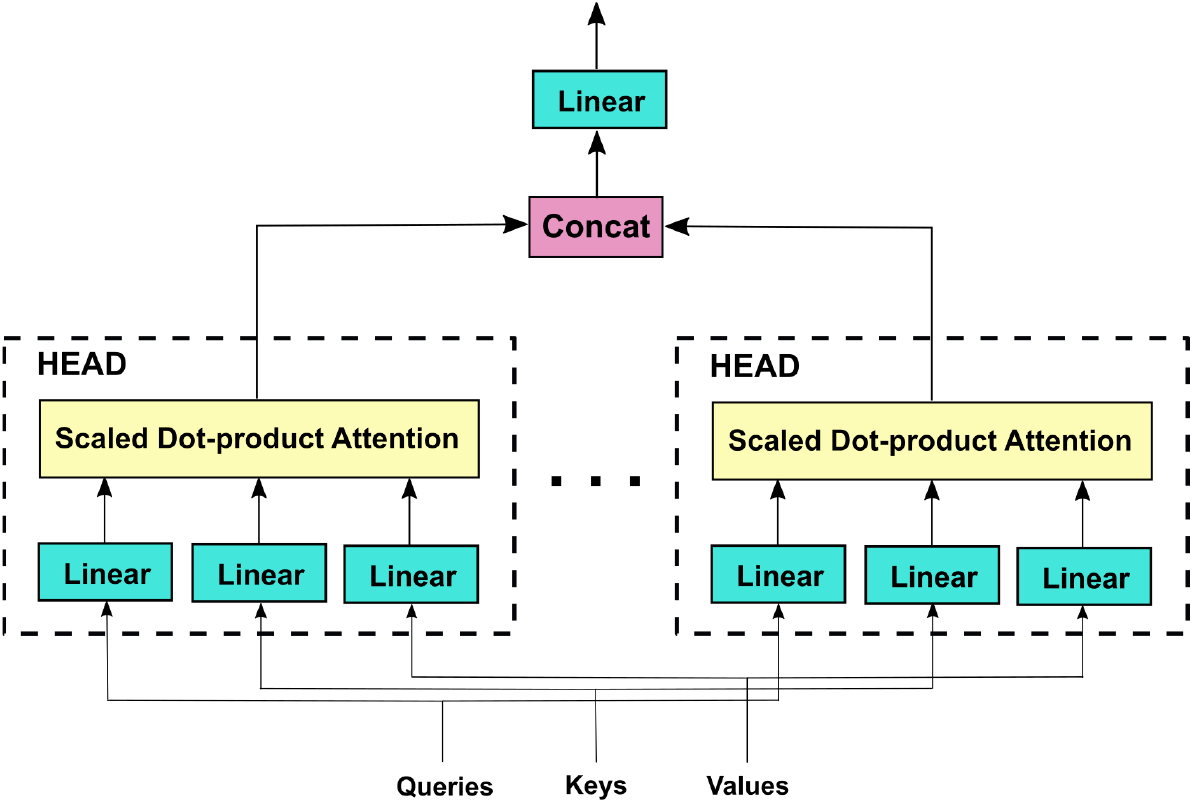
Multi-head Attention

In this study, the input queries, keys, and values of the multi-head attention block were duplicates of the convolution’s output after being positional encoded. Then, the three’s feature dimension would be equally split for *h* = 2 parallel attention heads, each had the same feature dimension *d*_*q*_ = *d*_*k*_ = *d*_*v*_ = *d*/ = 512/2 = 256. The results of each head were concatenated, thus the output still had the shape of (256, 157, 512).

### 3.3.2 Add & Normalization

The Add&Norm blocks also play a key role in the model. It uses residual connection [24] to add the inputs after dropout [30] of the Multi-head Attention block and the Position-wise Feed Forward Network to their respective outputs. Then the sums are normalized over the feature dimension by Layer Normalization [31], which standardizes the features in one training example, to facilitate faster training and better generalization.

### 3.3.3 Position-wise Feed Forward Network

This block consists of two fully connected linear transformations with a ReLU activation function [32] in between. It accepts 3-dimensional input tensors of shape (*batch size, sequence length, num features*), and only applies to the feature dimension. In other words, the block performs on each position in the recording length independently (position-wise). It outputs a tensor with the same shape as its input.

### 3.4 The output layer

After the Transformer Encoder, the model used a Dense layer with 13 hidden units with a softmax activation to generate the positive probabilities of 13 classes.

## 4. EXPERIMENTAL SETTINGS

In our experiments, we split the dataset into two subsets for training and testing, with a ratio of 9:1. With regard to hardware, we trained the models on an NVIDIA Tesla P100 GPU, repeatedly during 50 iterations (epochs). In each iteration, we split the training set into batches of 256 recordings each. To mitigate the undesired effect from the very high imbalance dataset, we employed oversampling [33] for the minority positive class of each abnormality. This served to increase the ratio of positives and negatives in each batch. In addition, the model parameters were initialized by a Xavier initializer [34], and optimized by Adam algorithm [35]. The learning rate of the Adam optimizer was set to 0.001 and was scheduled during training by cosine annealing warm restart [36], to quickly start the optimization process, then gradually decrease the learning rate to find the optimal solution. Finally, we used several regularization techniques to deal with overfitting, such as batch normalization [37], dropout [30], weight decay [38].

## 5. EVALUATION METRIC

The output layer of the model generates a positive probability for each abnormality. Then by defining a threshold, the probability can be used to classify its input into either positive or negative:

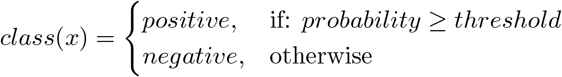

There are two types of error that a model can make in a classification problem: false positive and false negative. Precision and recall can be used to evaluate the two errors:

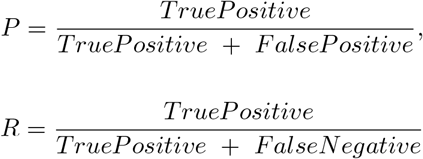

The formulae show that precision covers the falsepositive error, while recall takes false-negative cases into consideration. However, the two metrics cannot cover the other error type. In addition, the dataset was highly imbalanced, i.e., the ratio of positive and negative samples was very low for every abnormality, thus the model can be biased toward the majority negative class, having good precision and poor recall. Therefore, it is important to use a metric that is more balanced. One metric that can be used is *f*_1_-score, which is the harmonic mean of precision and recall, thus can better assess model performances.

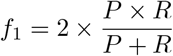

However, when using the precision, recall, and *f*_1_score, a classification threshold, 0.5 by default, must be specified in advance. It is better to evaluate models across the whole threshold range [0, 1] and enables us to find the optimal threshold value for each class. The area under receiver operating characteristics (AUROC) [39] and area under precision-recall curve (AUPRC) [40] are two such metrics. Nonetheless, [41] showed that the AUROC was likely to give misleading results when used on imbalanced datasets. Therefore, in this study, the AUPRC was used to evaluate our model. The threshold range [0, 1] was split into a large number of discrete values. For each value, we calculated the corresponding precision, recall, and *f*_1_-score, then plot a point that represents the precision at the *y*-axis and recall at the *x*-axis. Repeat the process for all threshold values results in a smooth curve, and the area under the curve is the AUPRC of the model, which also ranges from 0 to 1. The optimal threshold can be obtained by sorting the *f*_1_-score array.

## 6. RESULTS & DISCUSSIONS

In this work, we compared the performance of our proposed CNN-TE model with five different deep neural networks:

- VGG11 [42]: this model consisted of convolutional and fully connected layers, only capable of extracting spatial features.
- GRU [25] and LSTM [11]: the two popular RNNs for time-series tasks, only capable of extracting temporal information.
- CNN-GRU and CNN-LSTM: these stacked versions of convolutional and recurrent layers are able to extract both spatial and temporal features.

Table 2 shows the AUPRC of those models on 13 cardiac types in the test set. The CNN-TE outperformed other models in detecting all abnormalities, with AUPRC surpassed 0.8 on NSR, RBBB, STach, AF, MI, and LBBB. Visually, Figure 4 illustrates the precision-recall curves of all six models for those abnormalities. The curve of the CNN-TE was represented in blue, and it is clear that the area under the blue curve was larger than the areas under other curves. Meanwhile, CNN-GRU and CNN-LSTM had approximately the same performance. When using separately, LSTM was better than GRU, and the VGG11 performed poorly in most classes. Since the VGG11 was outperformed by GRU and LSTM, it could be inferred that temporal features in the ECG signals were much more important than spatial ones for classification. Additionally, stacking convolutional layers at the front significantly improved the RNNs’ performance, as shown in CNN-GRU and CNN-LSTM. Meanwhile, the CNN-TE model was not only capable of exploiting both types of features but also had intrinsic attention mechanisms, thus achieved the best results.

**Table 2.** Model performance comparison.

|  | VGG11 | GRU | LSTM | CNN-GRU | CNN-LSTM | CNN-TE |
| --- | --- | --- | --- | --- | --- | --- |
| NSR | 0.68 | 0.76 | 0.80 | 0.91 | 0.95 | 0.96 |
| RBBB | 0.69 | 0.74 | 0.79 | 0.87 | 0.88 | 0.89 |
| AF | 0.32 | 0.34 | 0.47 | 0.78 | 0.73 | 0.84 |
| STach | 0.62 | 0.23 | 0.29 | 0.57 | 0.73 | 0.83 |
| LBBB | 0.61 | 0.61 | 0.65 | 0.76 | 0.81 | 0.82 |
| MI | 0.25 | 0.38 | 0.49 | 0.73 | 0.76 | 0.80 |
| LAD | 0.39 | 0.50 | 0.60 | 0.73 | 0.77 | 0.79 |
| LAnFB | 0.18 | 0.41 | 0.45 | 0.71 | 0.74 | 0.78 |
| SB | 0.48 | 0.22 | 0.27 | 0.59 | 0.62 | 0.78 |
| IABV | 0.09 | 0.09 | 0.10 | 0.52 | 0.12 | 0.77 |
| abQRS | 0.12 | 0.32 | 0.41 | 0.68 | 0.66 | 0.70 |
| LVH | 0.19 | 0.28 | 0.34 | 0.58 | 0.63 | 0.69 |
| MIIs | 0.12 | 0.25 | 0.34 | 0.51 | 0.51 | 0.57 |

**Fig 4.**
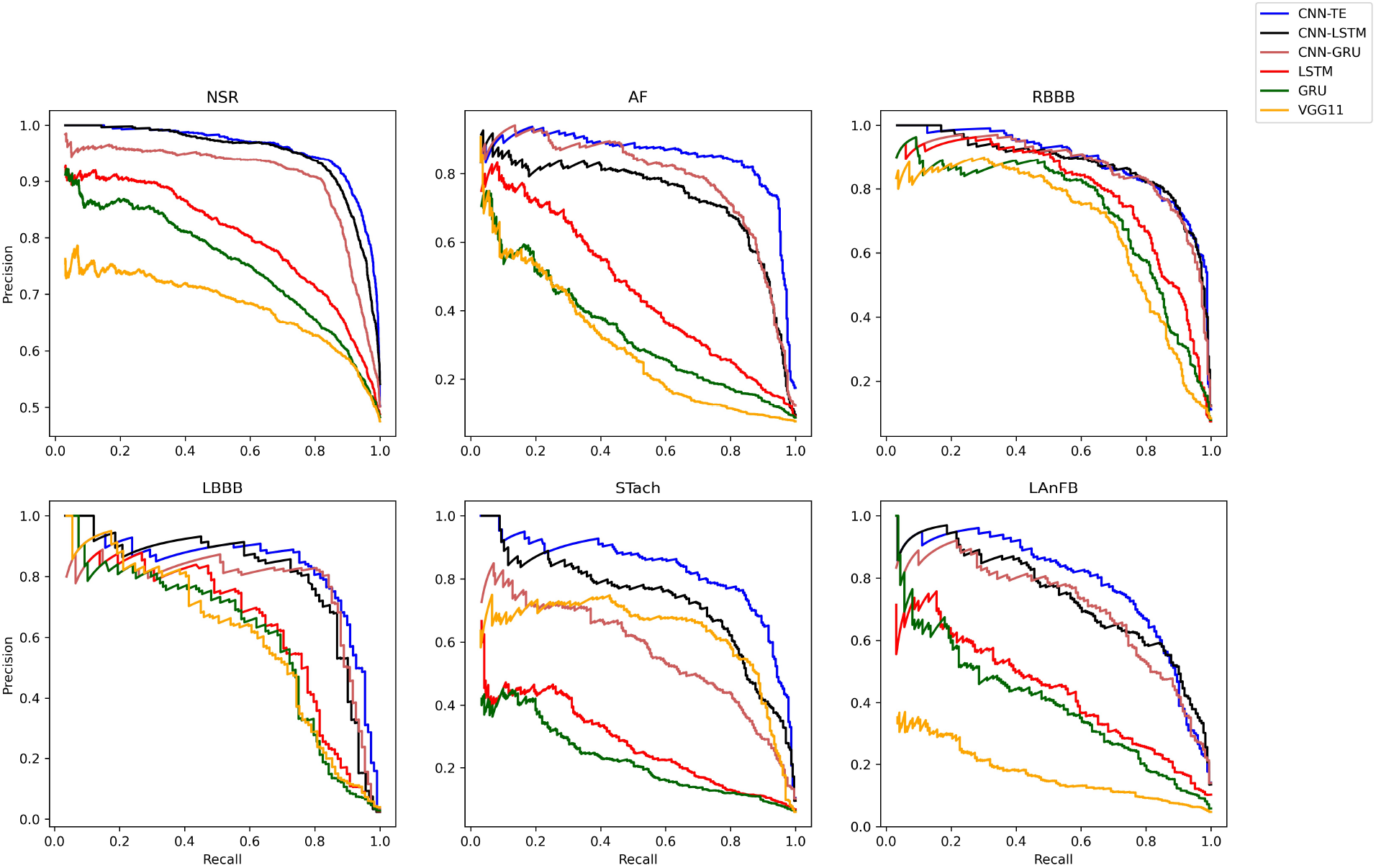
Precision-Recall curves of all six models for NSR, AF, RBBB, LBBB, STach, and LanFB

Our evaluation method allowed the optimal threshold that maximized the *f*_1_-score of a class to be obtained. Table 3 shows optimal classification threshold for each cardiac types, with the corresponding precision, recall and *f*_1_-score. All optimal thresholds were larger than the naive value of 0.5. which was expected for datasets that are heavily imbalanced towards the negative class. The Pearson product-moment correlation coefficient of -0.68 between the optimal thresholds and the percentage of positive samples indicated their strong negative correlation. In other words, the optimal threshold for a class tended to increase when there were fewer positive samples of that class.

**Table 3.** Optimal thresholds for the CNN-TE model.

| | Positive (%) | Optimal threshold | Precision | Recall | $f_1$-score |
| --- | --- | --- | --- | --- | --- |
| NSR | 48.84 | 0.68 | 0.91 | 0.89 | 0.90 |
| AF | 8.13 | 0.97 | 0.82 | 0.87 | 0.84 |
| RBBB | 7.25 | 0.84 | 0.81 | 0.83 | 0.82 |
| LBBB | 2.45 | 0.97 | 0.83 | 0.80 | 0.81 |
| STach | 5.62 | 0.88 | 0.76 | 0.84 | 0.80 |
| LAnFB | 4.25 | 0.89 | 0.73 | 0.76 | 0.74 |
| MI | 13.28 | 0.88 | 0.75 | 0.72 | 0.74 |
| LAD | 14.32 | 0.79 | 0.69 | 0.79 | 0.74 |
| SB | 5.55 | 0.97 | 0.71 | 0.75 | 0.73 |
| IABV | 5.63 | 0.90 | 0.71 | 0.72 | 0.71 |
| abQRS | 7.97 | 0.73 | 0.63 | 0.71 | 0.67 |
| LVH | 8.82 | 0.94 | 0.68 | 0.62 | 0.65 |
| MIIs | 6.02 | 0.84 | 0.53 | 0.67 | 0.59 |
| corr_coef | -0.68 |  |  |  |  |

## 7. CONCLUSION

In this study, we tried to tackle the multi-label classification problem in a medical context, detecting 13 cardiac types simultaneously from 12-lead ECG recordings. Specifically, we developed the combination of Convolution, Positional Encoder to explore spatial and temporal features in the signals, alongside a Transformer Encoder to utilize its multi-head attention mechanisms. We also took a proper evaluation method into consideration, by using a more comprehensive metric called AUPRC to assess model performance on the whole classification threshold range [0, 1]. Based on this method, we drew a comparison of how different deep learning models performed, obtained optimal threshold values for all cardiac types, then related it to the proportion of positive class. The results showed the effectiveness of our CNN-TE models that outperformed other networks. In addition, the strong negative correlation between the optimal threshold and the positive ratio indicated the significance of not relying on the naive threshold of 0.5 for medical classification tasks, which mostly involve heavily imbalanced datasets.

## Data Availability

All data produced are available online at the Physionet website.

